# Renal Involvement in Patients with COVID-19 Pneumonia and Outcomes After Stem Cell Nebulization

**DOI:** 10.1101/2020.12.16.20236877

**Authors:** Gina M. Torres Zambrano, Carlos A. Villegas Valverde, Antonio Bencomo Hernández, Lobna Abdel Hadi, Rene Antonio Rivero, Yendry Ventura Carmenate

**Author notes:** All authors participated on the critical revision of the manuscript for important intellectual content. Mailing address: Street #31, Area Muroor Road, Building # C43, Al Murjan Tower, Apartment 107, Abu Dhabi, UAE. **Abu Dhabi Stem Cells Center (ADSCC)** Ringgold ID: **580443**.

## Abstract

**Background:** The COVID-19 pandemic presented an unprecedented challenge to identify effective drugs for prevention and treatment.

**Objective:** To characterize acute renal injury (AKI) in patients with COVID-19 and their relation with clinical outcomes within the framework of the SENTAD COVID clinical trial at the Abu Dhabi Stem Cells Center.

**Methods:** Abu Dhabi Stem Cell Center (ADSCC) proposed a prospective clinical trial nebulization treatment with autologous stem cells (Non-Hematopoietic Peripheral Blood Stem Cells (NHPBSC)), at Abu Dhabi hospitals.

**Participants:** 20 treated patients were compared with 23 not treated patients. Both groups received COVID 19 standard treatment.

**Outcomes:** After the results were collected, this study was created to determine the impact of the disease on the renal function and the efficacy of the therapy on patient’s outcomes.

**Results:** One third of the critical patients studied suffered kidney failure. Patients in the treated group showed a favorable tendency to improve in contrast to those in the control group. Less patients from group A suffered from sepsis in comparison with the group B (25% vs 65%), HR=0.38, (95% Confidence Interval: 0.16 – 0.86), *p=0.0212. These results suggested a NNT=2.5. An improvement in lymphocyte count, CRP, and shorter hospital stay after treatment was evidenced, which led to less superinfection and sepsis in the treated group.

**Conclusions:** The proposed anti-inflammatory effect of the stem cells, offers a great promise for managing the illness, emerging as a crucial adjuvant tool in promoting healing and early recovery in severe COVID-19 infections and other supportive treatments.

**ARTICLE SUMMARY:** Our study had several strengths and limitation:

- It was a randomized trial.
- The treatment showed a positive result, providing evidence that this intervention is effective in routine practice.
- We found fewer complications related to prolonged hospital stay in the treated group.
- The is the small number of participants.
- It was carried out in 4 different hospitals, each with different criteria for the selection of the initial empirical antimicrobials, which can cause multiple resistant germs.

## INTRODUCTION

The COVID-19 pandemic caused by the novel severe acute respiratory syndrome coronavirus 2 (SARS-CoV-2) presented an unprecedented challenge to identify effective drugs for prevention and treatment. Many countries, including the UAE are taking every precautionary measure to slow and control the spread. (2) Society required the intervention of all the all the healthcare provider. Everyone got involved in different ways all over the world, especially as physicians, we had to support the work at the intensive care unit (ICU), no matter if we were urologist.(3)

In order to assist with local research efforts to discover therapies for COVID-19, the research team at Abu Dhabi Stem Cell Centre (ADSCC) proposed a clinical trial with the use of autologous stem cells, called Non-Hematopoietic Peripheral Blood Stem Cells (NHPBSC), taking into account the expected safety profile of these biological products, and its immunomodulatory capacities. The SENTAD COVID study began in April 2020; this treatment Study was adaptive, prospective, involving hospitalized adult patients with confirmed COVID-19 infection during the outbreak in Abu Dhabi, 2020.(4) The potential breakthrough coronavirus treatment developed at ADSCC has shown promising results.

The virus and comorbidities related were seen to affect the urinary tract and kidney function, being able to become an important factor of morbidity and mortality. Zou et al., (5,6) studied the expression of angiotensin-converting enzyme (ACE2) positive cells, targets for the SARS-CoV-2 infection,(6,7) and determined organs at high risk of viral invasion included the kidneys (they found ACE2 positive cells at the proximal convoluted tubules in about 4%). This could be one of the factors that caused acute kidney injuries in 0.1–29% of the patients with COVID-19.

Another important factor is the sepsis or septic shock resulting in cytokine storm syndrome (induced due to the high levels of cytokines, and consequently, numerous adverse reactions in the human body are observed) or by the immune-mediated kidney damage,(6,8) causing high mortality rates (60–90%).(8) Given the potential of the stem cells in sepsis and clinical evolution of chronic conditions, with the stimulation by humoral factors, strategies such as stem cell-based therapy are being proposed to regulate inflammation, prevent or mitigate this cytokine storm through their immunomodulatory capacity.(9)

Tools like neutrophil-lymphocyte ratio (NLR), were being used as a prognostic biomarker in urological diseases like kidney and bladder cancers,(10,11) during the pandemic, several authors used this ratio as an independent indicator of short- and long-term mortality, which does not require additional costs and is rapidly accessible.(12,13) A recent meta-analysis noted that 35–75% of COVID patients admitted at the ICU developed lymphopenia, which was a more frequent feature of patients who died of the disease.(14,15) In the analysis of 67 COVID-19 patients from Singapore, a low lymphocyte count was reported, being therefore predictive for admission to the intensive care unit (ICU).(14,16) In this article we analysed the repercussion of the disease in the kidney function and the stem cell treatment given with the outcomes.

## OBJECTIVE

To characterize acute renal injury (AKI) in patients with COVID-19 and their relation with clinical outcomes within the framework of the SENTAD COVID clinical trial at the Abu Dhabi Stem Cells Center.

## METHODS AND PATIENT INVOLVEMENT

The SENTAD COVID Study was conducted at the Emirate of Abu Dhabi in 4 different hospitals: Sheikh Khalifa Medical City, Al Rahba Hospital, Al Mafraq Hospital and Al Ain Hospital. This was a prospective and analytical study implemented to determine the impact of the disease on the renal function and the efficiency of the therapy on patient outcomes within the framework of the SENTAD COVID clinical trial at ADSCC.(4) A comparison between critical patients in the group A (Treated) and group B (Controls) was done. The Inclusion criteria is as follows: Aged ≥ 18 years, laboratory confirmation of COVID-19, interstitial lung change judged by computed tomography, hospitalized and symptomatic patients, referring one or more symptoms (fever, cough, or shortness of breath, tiredness, runny nose, headache, sore throat, chills, muscle pain, or new loss of taste or smell), ability to comply with test requirements and blood collection and agrees to participate in the study.

The Exclusion criteria is as follows: Pediatric patients (aged < 18 years), diagnosis of any kind of shock, organ transplants in the past 3 months, patients receiving immunosuppressive therapy, diagnostic of Hepatitis B Virus (HBV) infection or Human Immunodeficiency Virus (HIV) infection or Acquired Immunodeficiency Syndrome (AIDS), current diagnosis of cancer or History of malignancies in the past 5 years, pregnant or lactating women, patients who had participated in other clinical trials in the past 3 months, inability to comply with test requirements and blood collection or inability to provide informed consent.

In the main study, the ordinal scale created by the WHO committee for COVID-19, for clinical involvement overtime was used in order to determine the severity,(17) then, the critical patients, having scores between 5 to 7, were selected to rule out the renal involvement: Patients with non-invasive ventilation or high-flow oxygen were assigned a score of 5, intubation and mechanical ventilation score of 6, mechanical ventilation + additional organ support: ECMO, CRRT or vasopressors score of 7, and finally death score of 8.

The analysis of the demographic variables from both arms, including gender, age, BMI, and comorbidities, vital signs, and biochemical studies performed within 24 h of inclusion in the trial. Furthermore, a record of plasma-based and serum-based biomarkers concentrations, including high-sensitivity C-reactive protein, D-dimer, and interleukin-6 (IL-6) was carry out. An analysis of the creatinine, thrombocytosis and neutrophil/lymphocyte ratio were done to calculate the independent indicator of mortality. Lymphopenia was defined as an absolute lymphocyte count <1000 × 106 /L. NLR abnormal value cut point used was ≥3.(12,13)

Both groups received COVID 19 standard treatment defined as “UAE National Guidelines for Clinical Management and Treatment of COVID 19, v.2.0” as per the Department of Health (DOH).(18) For patients at group A, a stem cell nebulization with Autologous non hematopoietic peripheral blood stem cells (NHPBSC, a cocktail of V-cells and grow factors, named UAECell 19), obtained through phlebotomy of 300cc and processed at the ADSCC laboratory, and were characterized by flow cytometry and microscopy (Fig.1). Follow up of both groups were done until they were discharged, measuring controlled blood analysis when it was possible. Subsequently, collection of the length of hospital stay and the interval between the intervention (date from treatment in the case of group A, or time from inclusion in the cases of group B) and discharge date.

**Figure 1.**
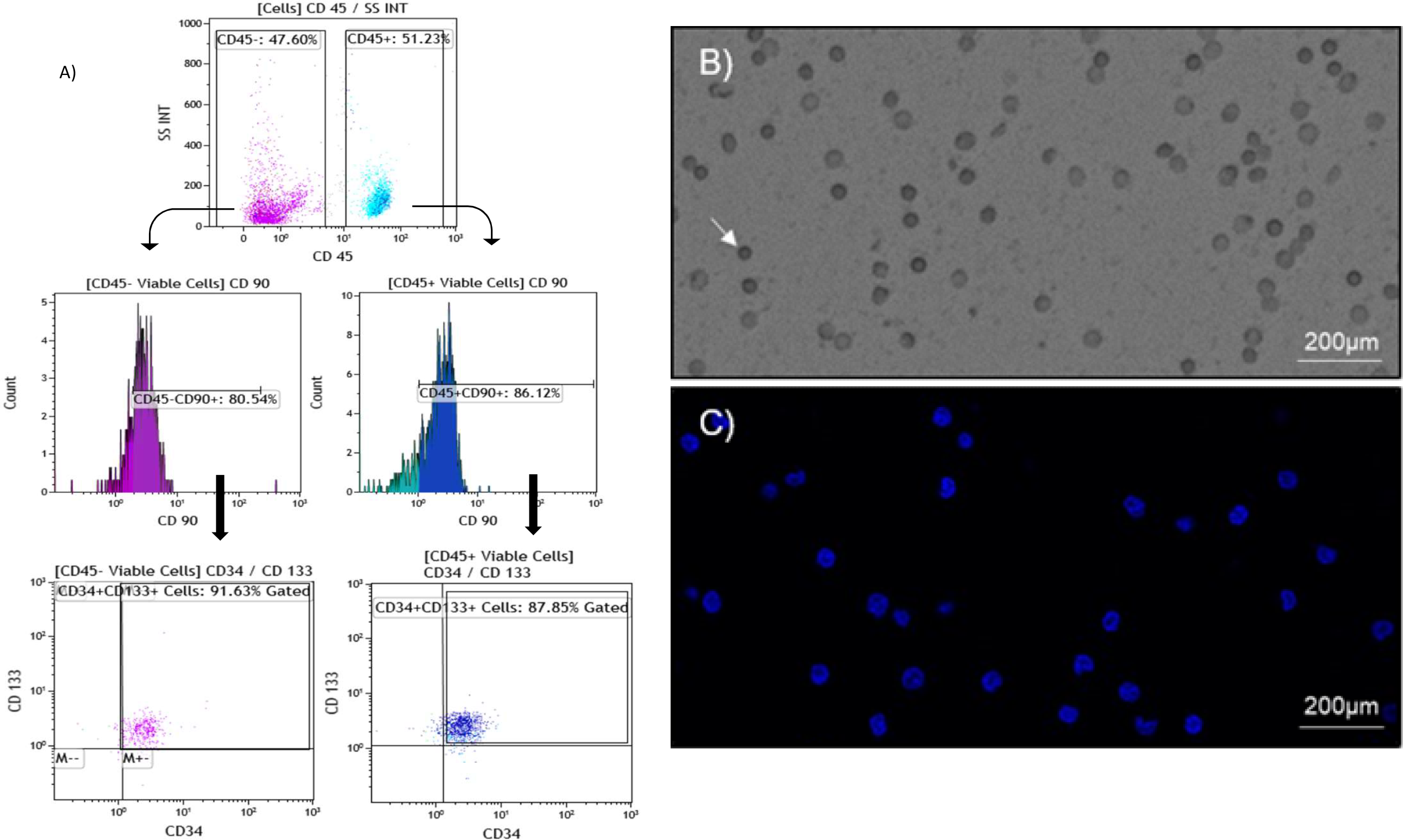
Characterization cytomorphologic of autologous hematopoietic and non hematopoietic peripheral blood stem cells (NHPBSC). A) Flow cytometric assessment of NHPBSC with a manual and hierarchical gating strategy. Two subpopulations are described according to the expression of CD45 and characterized by the co-expression of CD133+/CD90+/CD34+. Representative images taken by Lionheart TM FX Automated Microscope of NHPBSC population, with the presence of Erythrocytes contamination, after 24 h of isolation from COVID-19 infected peripheral blood samples. B) Bright field Microscopic image of NHPBSC morphology, indicated by white arrow. C) DAPI (5µg/ml) DNA staining showing the nucleus. Bars in B) and C) indicate 200µm (original magnification, 10x).

### Patient and Public Involvement

the trial came about in response to the COVID - 19 pandemic and the need for innovative treatments. The patients were recruited from the governmental hospitals, making sure they full fill the inclusion criteria and signed the informed consent, they were not involved in setting the research question, but they were they intimately involved in the implementation of the intervention. Nowadays, they are being informed about the results of the trial.

### Statistical analysis

a non-normal distribution of the variables was found, so non-parametric statistical methods were used. A proportions comparison test (Chi-Square) for the BMI categories (19) and comorbidities, and U-Mann-Whitney for the dates intervals and laboratory parameters. The Hazard Ratio (HR), 95% Confidence Interval, Number Needed to Treat (NNT) was calculated for the sepsis complication. In addition, follow-up laboratory tests were compared up to 25 days after therapy, using a Wilcoxon test in group B.

## RESULTS

From the main study, a total of 139 patients were divided in two groups for the comparison:

1. Group A (Experimental arm) COVID 19 standard care plus nebulization with PBNHESC (n=69)
2. Group B (No intervention arm) COVID 19 standard care (n=70).

Following the seven-category ordinal scale for clinical involvement, 44 patients (31%) were assigned as critical. A total of 20 patients (29%) from the group A and 24 patients (34%) from the group B where compared, all the patients were male. The ages of group A range from 32 to 71 years old (mean 49), and from 26 to 67 years old in group B (mean 48.03). The distribution of the BMI was between 17 to 33.42 (mean 26.51) in the group A, and between 22.7 to 45.71 (mean 30.01) in the group B. No significant differences were found between the two groups on age (p=0.75) and BMI (p=0.0943). Regarding the comorbidities, Diabetes Mellitus and hypertension were the most commons diseases in both groups, without differences between them (Table 1).

**Table 1.** Clinics characteristics of the patients.

|  | Group A |  | Group B |  | p* |
| --- | --- | --- | --- | --- | --- |
|  | n | % | n | % |  |
| BMI: |  |  |  |  |  |
| Overweight (25-29) | 9 | 45% | 10 | 42% | 0.8433 |
| Mild obesity (30-34) | 3 | 15% | 6 | 25% | 0.4182 |
| Moderate obesity (35-40) | 0 | 0% | 2 | 8% | 0.2010 |
| Morbid obesity (>40) | 0 | 0% | 2 | 8% | 0.2010 |
| Comorbidities |  |  |  |  |  |
| Smoker | 1 | 5% | 0 | 0% | 0.2733 |
| DM | 9 | 45% | 7 | 29% | 0.2772 |
| HTN | 5 | 25% | 6 | 25% | 1 |
| DL | 3 | 15% | 1 | 4% | 0.2095 |
| Cardiac diseases | 1 | 5% | 1 | 4% | 0.8742 |
| Respiratory diseases | 2 | 10% | 1 | 4% | 0.4342 |
| Score |  |  |  |  |  |
| 5 | 3 | 15% | 7 | 29% | 0.2747 |
| 6 | 2 | 10% | 3 | 13% | 0.7600 |
| 7 | 15 | 75% | 15 | 63% | 0.3991 |

BMI: Body Mass Index. DM: Diabetes Mellitus. HTN: Hypertension. DL: Dyslipidemia. \* p: proportions comparison test.

Creatinine concentrations, total leucocytes, IL-6, high-sensitivity C-reactive protein and D-dimer were elevated in most patients, without significant differences between both groups (Table 2).

**Table 2.** Laboratory parameters distribution.

| Parameters | Group A n=20 |  | Group B n=24 |  | p |
| --- | --- | --- | --- | --- | --- |
|  | Median | Range (2.5-97.5) | Median | Range (2.5-97.5) |  |
| IL6 | 450.6 | (20.33 - 10016.0) | 233 | (23.7 - 5000.0) | 0.7517 |
| D-dimer (mg/mL) | 1.78 | (0.58 - 8.70) | 1.45 | (0.310 - 35.20) | 0.9998 |
| CRP (mg/L) | 209.8 | (6.5 - 444.3) | 155.1 | (0.8 - 539.7) | 0.2211 |
| WBC | 9500 | (800 - 23100) | 9500 | (2900 - 41800) | 0.8613 |
| Neutrophils ($\times 10^6/L$) | 8700 | (4200 - 21000) | 6770 | (2280 - 14900) | 0.1609 |
| Lymphocytes ($\times 10^6/L$) | 800 | (400 - 1900) | 1190 | (570 - 2470) | *0.0139 |
| Platelet $\times 10^3/\mu L$ | 271.5 | (142 - 599) | 238 | (15.10 - 481) | 0.3299 |
| Creatinine (mg/L) | 1.17 | (0.58 - 6.11) | 1.14 | (0.57 – 7.93) | 0.9543 |

L6: interleukin 6; CRP: C-reactive protein; WBC: white blood cells; CRP: C-reactive protein; p: U-Mann-Whitney; Range: percentile 2.5-97.5. \*: significant.

Lymphocytopaenia was common in both groups, but more considerable in group A (range 400 - 1900) compared with group B (range 570 - 2470), showing a significant difference (p= *0.0139), favoring group B. The NLR ≥3, gave a higher risk of mortality in patients in group A (100%), compared to patients in group B (79%), before the intervention (p=0.0313) (Table 3). Moreover, after the intervention in group A the NLR dropped from 20 patients (100%) to 15 patients (75%) with statistical significance (p= 0.0183).

**Table 3.** Distribution of neutrophil-lymphocyte ratio (NLR) in the patients before and after intervention.

| Time of assessment | Group A | Group B | p |
| --- | --- | --- | --- |
| Recruitment | 100% | 79% | *0.0313 |
| 25 day after | 75% | 79% | 0.7556 |
| p | *0.0183 | 1 |  |

p: Comparison of Proportions. \*: significant.

The patients from group A received two nebulization of 10cc with UAECell 19, in two consecutive days (at least 22 hours between the therapy). The comparison between groups after the intervention (Table 4), showed a shorter hospital stay in group A with a mean of 27.4 days, while in group B it was 41.6 days, with a maximum of only 45 days in the group A, compared with 126 days in the group B. Also, the interval from the intervention day until the discharge, group A had a maximum of 43 days compared with group B with 125 days. In group B there were 6 deaths while in the group A there were only 4 (p: 0.4535). A quarter of the patients of the group A required continuous renal replacement therapy for AKI with hemodialysis, compared with 35% of the patients on group B.

**Table 4.**
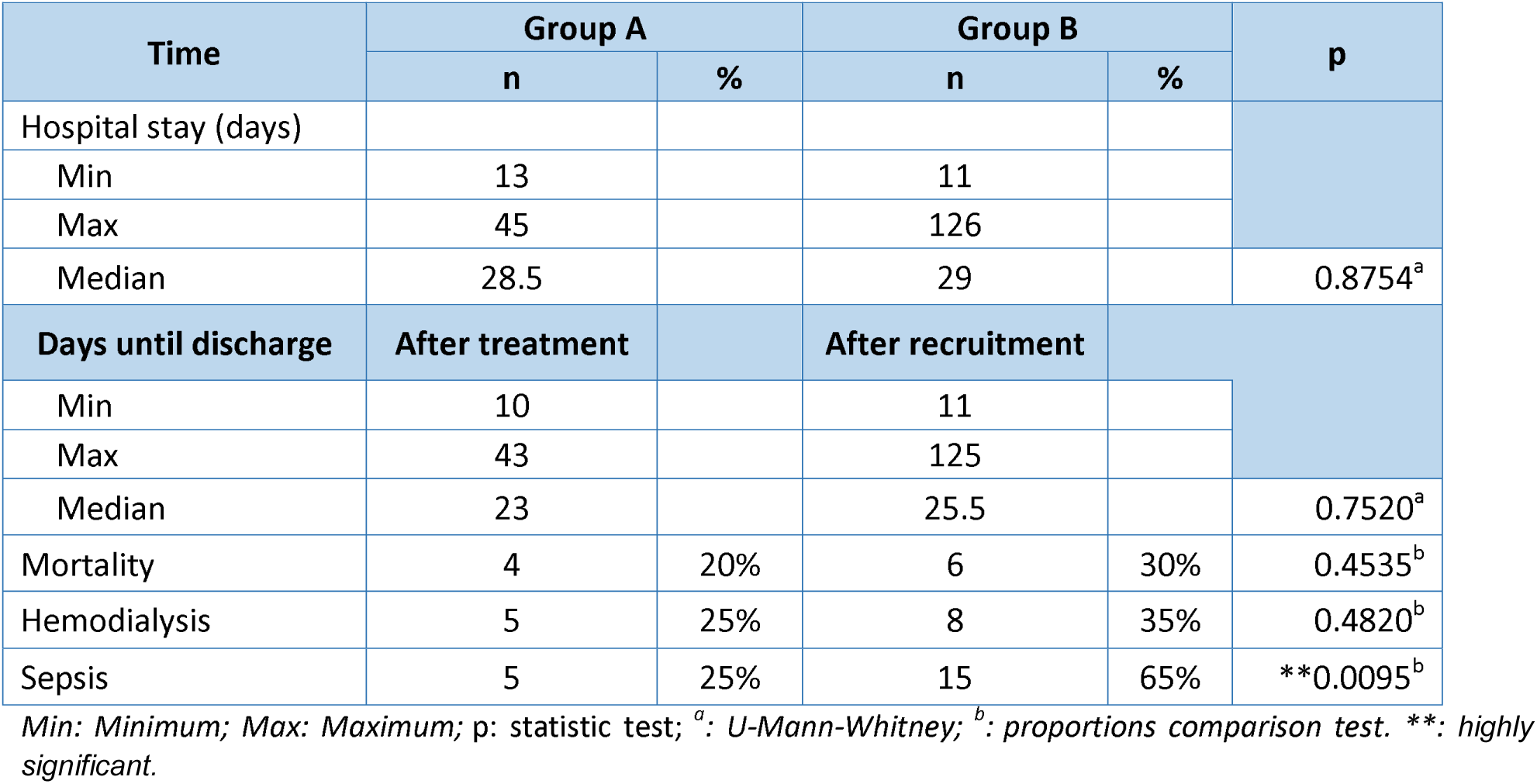
Clinical outcome characterizations.

A lower proportion of patients from group A suffered from sepsis in comparison with group B (25% vs 65%), HR=0.38, (95% Confidence Interval: 0.16 – 0.86), *p=0.0212. These results suggested a NNT=2.5 (table 4). The pathogens causing sepsis in group A vs. group B were: *Candida albicans* (5% vs 22%), *Streptococcus pneumonia* (10% vs 0%), *Pseudomonas aeruginosa* (5% vs 9%), *Klebsiella pneumoniae* (0% vs 17%), *Enterobacter aerogenes* (0% vs 4%) and *Staphylococcus aureus* (5% vs 4%).

In the group A, the creatinine, WBC, neutrophils and platelet count, did not show important variations during the interval of study, but the CRP and lymphocytes count were drastically down after the treatment (Table 5).

**Table 5.**
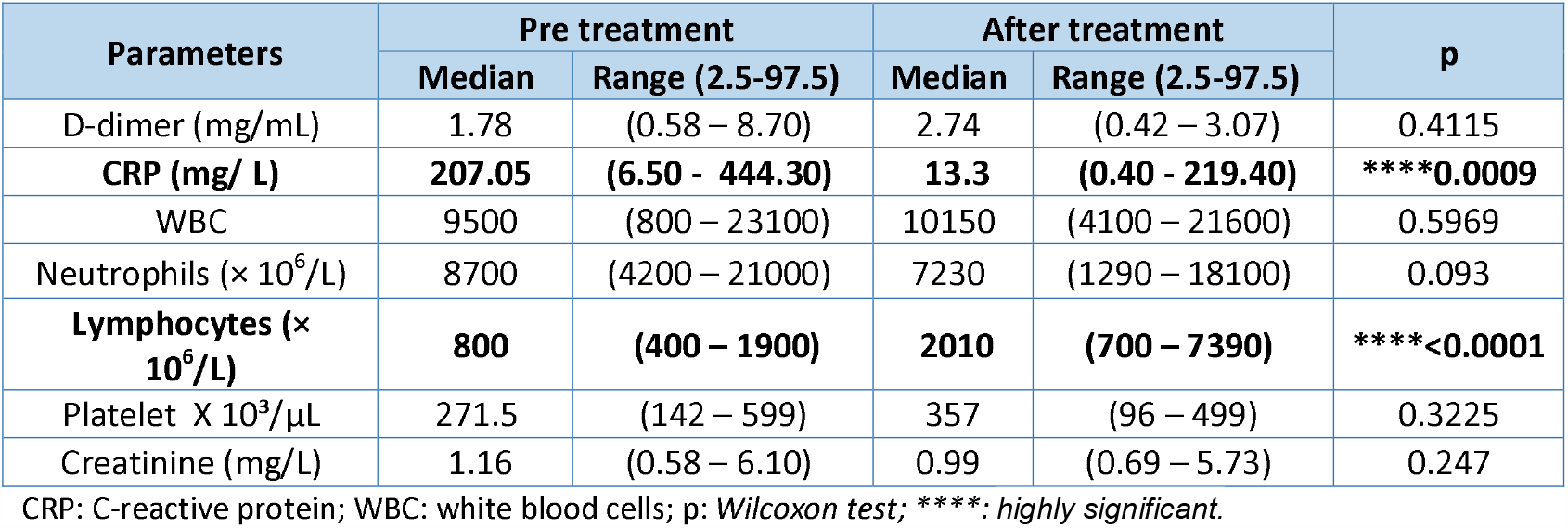
Laboratory findings pre and after treatment in Group A.

| Parameters | Pre treatment |  | After treatment |  | p |
| --- | --- | --- | --- | --- | --- |
|  | Median | Range (2.5-97.5) | Median | Range (2.5-97.5) |  |
| D-dimer (mg/mL) | 1.78 | (0.58 – 8.70) | 2.74 | (0.42 – 3.07) | 0.4115 |
| **CRP (mg/ L)** | **207.05** | **(6.50 - 444.30)** | **13.3** | **(0.40 - 219.40)** | **\*\*\*\*0.0009** |
| WBC | 9500 | (800 – 23100) | 10150 | (4100 – 21600) | 0.5969 |
| Neutrophils (× 10<sup>6</sup>/L) | 8700 | (4200 – 21000) | 7230 | (1290 – 18100) | 0.093 |
| **Lymphocytes (× 10<sup>6</sup>/L)** | **800** | **(400 – 1900)** | **2010** | **(700 – 7390)** | **\*\*\*\*<0.0001** |
| Platelet X 10<sup>3</sup>/μL | 271.5 | (142 – 599) | 357 | (96 – 499) | 0.3225 |
| Creatinine (mg/L) | 1.16 | (0.58 – 6.10) | 0.99 | (0.69 – 5.73) | 0.247 |

CRP: C-reactive protein; WBC: white blood cells; p: Wilcoxon test; \*\*\*\*: highly significant.

## DISCUSSION

The data analyzed of this study, showed a higher incidence of COVID-19 among males, especially those admitted in the ICU (no female with critical scores were found during the study period). The early experience from China confirmed the male predominance in the incidence of this disease (58%) compared to female, it has also been found that patients with diabetes, hypertension, coronary heart disease, chronic obstructive pulmonary disease, cerebrovascular disease, and kidney disease exhibit worse clinical outcomes when are infected with COVID-19.(20,21) Similar to data reported elsewhere, the critical patients included in the study were found to have cardiopulmonary comorbidities, diabetes mellitus, hypertension and higher concentrations of D-dimer as independent risk factors for poor outcomes.(20,21)

Recent evidence shows that AKI is common in COVID-19 critical ill patients. Accordingly, one study from Wuhan, China, found in 52 critically ill patients admitted in the ICU that AKI was the most common extra-pulmonary complication, present in 15 patients (29%), more common than cardiac injury (23%), and liver dysfunction (23%). Of all AKI patients, eight (25%) needed continuous renal replacement therapy and 12 (80%) death with a median duration from ICU admission to death of 7 days (IQR 3-11).(20,22) All this together, suggest that kidney abnormalities are more common than expected and are a fatal complication associated with higher mortality.

In our study, the hospital stays after inclusion, mortality and specially the sepsis were less in group A contrast with group B. Despite the fact that the median hospital stay after the intervention was not significant between the groups, the range is much shorter in group A patients than in group B, showing a greater homogeneity in group A, probably related with the treatment (that facilitates the prediction of disease behavior), which was around 32 days, while group B was around 115 days due to the statistical dispersion of the data of this last group.

The lower incidence of sepsis in group A can be explained by this phenomenon, since the shorter hospital stay is a protective factor against nosocomial infections,(23) and it was found that around 3 patients are required to be treated to have this protective effect with the stem cell nebulization (NNT=2.5). An attenuation of bacterial sepsis mediated by stem cells had been described, via several mechanisms, such as improving the phagocytic ability, secreting anti-microbial peptides,(9,24) and increasing bacterial clearance.(9,25)

Likewise, prognostic biomarkers improved after receiving the treatment, such as CRP and lymphocytes count, and consequently the NLR. In our study, all critical ill patients had elevated acute phase reactants before the intervention, but the group A patients were found to have a higher risk of mortality, due to the lynphopenia and high NLR,(13) related to the severity of the disease described in this type of patient, while in group B there were no changes in the follow up of these biomarkers.

There is a polemic discussion among the clinicians regarding the pathophysiology of the kidney damage. It was demonstrated the virus presence in tubular epithelial cells by immunohistochemistry and in situ hybridization.(20,26) but other authors suggested that AKI in COVID-19 patients could be the result of cytokine release syndrome,(27) rather than active viral replication in the kidney. It is well known that stem cells have the potential to differentiate into lung cells among others, reprogramming the immune response to reduce destructive inflammatory elements and directly replace damaged cells and tissues.(9) Furthermore, the growth factor cocktail has apocrine effects on the modulation of the cytokine storm, not only in the lungs but throughout the circulatory system, explaining this phenomenon found in our study, which was evidenced by a decrease in acute phase reactants. It is almost certain that if we had the chance to treat those patients earlier, we would be able to avoid the AKI and hemodialysis requirement, since the immunomodulation with the therapy would suppress the cytokine storm. Additionally, because this therapy is an autologous treatment obtained after the blood donation of the same patient, there are no immunological reactions to this preparation, with rare adverse effects (fainting and dizziness may occur linked with the blood collection).

## CONCLUSION

In this study about one third of the critically ill patients had kidney failure. Patients in the treated group showed a better tendency to improve than those in the control group. COVID-19 affects kidney pathophysiology in different ways, therefore, a rational approach to possible therapeutic strategies due to the proposed immunomodulatory effect of the stem cells, offers a great promise for disease management, emerging as a crucial adjuvant tool in promoting healing and early recovery in severe COVID-19 infections and other supportive treatments.

## Data Availability

Extra data is available by emailing.

## ACKNOWLEDGMENTS

Acknowledgment to the government of the United Arab Emirates and especially to the Abu Dhabi Health Services Company SEHA and all its healthcare workers for the contribution and support to this study.

## ETHICS

The study was approved by the Emirates Institutional Review Board for COVID-19 Research (Ref. ID: DOH/CVDC/2020/1172). Study participants provided written informed consent per the Helsinki Declaration.(1) The consent document described the importance of participation and explained the study’s characteristics and possible risks and benefits. All data were kept confidential and participant identity was delinked. The selection of diagnostic tools followed the ethical principles of maximum benefit and non-maleficence. This work was supported by ADSCC. This research had not received any other specific grant from any funding agency in the public, commercial or not-for-profit sectors. Award/Grant number is not applicable. None of the contributing authors have any conflicts of interest. Extra data is available by emailing.

